# Targeted long-read sequencing as a single assay improves diagnosis of spastic-ataxia disorders

**DOI:** 10.1101/2024.09.04.24312938

**Authors:** Laura Ivete Rudaks, Igor Stevanovski, Dennis Yeow, Andre L. M. Reis, Sanjog R. Chintalaphani, Pak Leng Cheong, Hasindu Gamaarachchi, Lisa Worgan, Kate Ahmad, Michael Hayes, Andrew Hannaford, Samuel Kim, Victor S. C. Fung, Michael Halmagyi, Andrew Martin, David Manser, Michel Tchan, Karl Ng, Marina L. Kennerson, Ira W. Deveson, Kishore Raj Kumar

**Affiliations:** Molecular Medicine Laboratory and Neurology Department, Concord Repatriation General Hospital, Concord, 2137, Australia; Faculty of Medicine and Health, University of Sydney, Camperdown, 2050, Australia; Genomic and Inherited Disease Program, The Garvan Institute of Medical Research, Darlinghurst, 2010, Australia; Clinical Genetics Unit, Royal North Shore Hospital, St Leonards, 2065, Australia; Centre for Population Genomics, The Garvan Institute of Medical Research, Darlinghurst, 2010, Australia; Neurodegenerative Service, Prince of Wales Hospital, Randwick, 2031, Australia; Neuroscience Research Australia, Randwick, 2031, Australia; Faculty of Medicine, University of New South Wales, Kensington, 2052, Australia; School of Computer Science and Engineering, University of New South Wales, Kensington, 2052, Australia; Clinical Genetics Service, Royal Prince Alfred Hospital, Camperdown, 2050, Australia; Neurology Department, Royal North Shore Hospital, St Leonards, 2065, Australia; Movement Disorders Unit, Neurology Department, Westmead Hospital, Westmead, 2145, Australia; Neurology Department, Royal Prince Alfred Hospital, Camperdown, 2050, Australia; Department of Genetic Medicine, Westmead, 2145, Australia; The Northcott Neuroscience Laboratory, ANZAC Research Institute, Sydney Local Health District, Concord, 2137, Australia; School of Clinical Medicine, Faculty of Medicine and Health, St Vincent’s Healthcare Clinical Campus, University of New South Wales, Darlinghurst, 2010, Australia

**Keywords:** Nanopore sequencing, hereditary cerebellar ataxia, spinocerebellar ataxia, hereditary spastic paraplegia

## Abstract

The hereditary spastic-ataxia spectrum disorders are a group of rare disabling neurological diseases. The genetic testing process is complex, and often requires multiple different assays to evaluate the many potential causative genes and variant types, including short tandem repeat expansions, single nucleotide variants, insertions/deletions, structural variants and copy number variants. This can be a protracted process and, even after all avenues are exhausted, many individuals do not receive a genetic diagnosis.

Aiming to streamline and improve this process, we developed a targeted long-read sequencing strategy with capacity to characterise genetic variation of all types and sizes within 469 disease-associated genes, in a single assay. We applied this to a cohort of 34 individuals with genetically undiagnosed spastic-ataxia spectrum disorders. An additional five individuals with a known genetic diagnosis were included as positive controls.

We identified causative pathogenic variants that would be sufficient for genetic diagnosis in 14/34 (41%) unsolved participants. The success rate was 5/11 (45%) in those who were naïve to genetic testing and 9/23 (39%) in those who were undiagnosed after prior genetic testing, completed on a clinical basis. Short tandem repeat expansions in *FGF14* were the most common cause, present in 7/34 (21%). Two individuals (2/34, 6%) had biallelic pathogenic short tandem repeat expansions in *RFC1* and one individual had a monoallelic pathogenic short tandem repeat expansion in *ATXN8OS*/*ATXN8*. Causative pathogenic sequence variants other than short tandem repeat expansions were found in four individuals, including a heterozygous missense variant in *VCP*, a heterozygous in-frame deletion in *STUB1*, a homozygous splicing variant in *ANO10*, and compound heterozygous missense and nonsense variants in *SPG7*. In addition to these solved cases, a pathogenic or likely-pathogenic variant with uncertain clinical implications was identified in a further three individuals, including a single individual who was found to have a short tandem repeat expansion in *BEAN1* in addition to biallelic expansions in *FGF14* within the range of 200-249 repeats.

Our results demonstrate the utility of targeted long-read sequencing in the genetic evaluation of patients with spastic-ataxia spectrum disorders, highlighting both the capacity to increase overall diagnostic yield and to streamline the testing pathway by capturing all known genetic causes in a single assay.

## Introduction

The hereditary spastic-ataxia spectrum disorders are a group of rare, disabling neurologic conditions encompassing the phenotypic spectrum from pure ataxia, mixed spastic-ataxia to spastic paraparesis^1^ with combined global prevalence estimated at 9.8/100,000^2^. Although historically, hereditary cerebellar ataxia (HCA) and hereditary spastic paraplegia (HSP) have been classified separately on a clinicogenetic basis, with the discovery of an increasing number of genes causing overlapping phenotypes and shared disease mechanisms, it has been proposed that these disorders be considered as part of a spastic-ataxia spectrum disorder^3^. There are numerous causative genes and variant types, which have typically necessitated the use of a combination of different genetic testing methods for identification. This includes Southern blot or repeat-primed PCR for short tandem repeat (STR) expansions; next-generation sequencing (NGS) (including targeted gene panels, whole exome sequencing [WES] or whole genome sequencing [WGS]) to evaluate conventional variants (single nucleotide variants [SNVs], insertions/deletions [indels] and structural variants [SVs]); and subsequent consideration for additional tests, such as multiplex ligation-dependent probe amplification (MLPA) for larger copy number variants (CNVs) that may not be detected by NGS^4–7^. However, despite advances in genetic testing methods and broader access to NGS, up to 71% of individuals with HCA and 45-50% of individuals with a HSP phenotype currently do not receive a genetic diagnosis^1,6,8–12^.

There are several potential contributors to this ‘diagnostic gap’. STR expansions, evaluated by repeat-primed PCR or Southern blot, require specific primers/probes for each gene^5^. Typically, only the most common STR expansions are evaluated as part of a ‘SCA panel’, the content of which may vary between sites, whilst other less common or recently described STR expansions may be overlooked. Furthermore, potentially relevant genes may be omitted from targeted testing of a narrow phenotype-specific gene set, which is particularly relevant in cases with less ‘classic’ or atypical phenotypic features. Certain genetic variants are challenging to detect with NGS, such as SVs or any variant within a repetitive genomic region^13^. Targeted gene sequencing and WES are unable to identify deep intronic variants^14^. Short-read NGS is also generally unsuitable for phasing variants onto maternal vs paternal alleles, for example, to establish compound heterozygosity in the absence of parental sequencing data^13^. There may also be issues with access or availability of genetic testing, especially for more recently described STR expansion disorders. For instance, presently in Australia, clinically accredited testing for STR expansions in *FGF14* or *RFC1*, which cause late-onset ataxia^4^, is not available locally.

Long-read sequencing (LRS) is an emerging group of genomic technologies, which overcome several limitations posed by earlier methods, including NGS^13^. LRS provides improved detection of STR expansions, SVs and CNVs; ability to resolve repetitive regions and homologous gene families or gene-pseudogene pairs; variant phasing without parental sequencing data; and DNA methylation profiling at no additional cost^13^. LRS is particularly advantageous for genotyping STR expansions, where it can clearly identify expansion size, sequence, methylation state and zygosity, even for large, complex or GC/AT-rich repeats. These capabilities are critical for evaluating genes where different STR motif conformations can affect pathogenicity; for example, 12 different *RFC1* STR expansion motifs have been identified, some of which are pathogenic and others benign^4,15^. LRS also allows identification of repeat interruptions or flanking genetic variation that can modify penetrance and prognosis^16,17^.

The adaptive sampling or ‘ReadUntil’ functionality on LRS instruments from Oxford Nanopore Technologies (ONT) enables selective sequencing of specific genes using genetic coordinates provided programmatically and without the need for additional laboratory processes for target enrichment^18^. Targeted LRS can be used to evaluate a large number of STRs in a single test and has demonstrated success in concurrent testing of a set of 37 genes associated with neurologic disease^16^ as well as concurrent evaluation of 10 STR loci associated with ataxia^15^. Although these studies establish the analytical validity of this approach for detection of STR expansions, they did not examine non-STR causative variants for ataxia/neurologic disease, nor assess the capacity of LRS to solve otherwise undiagnosed patients.

Given the capacity to evaluate a broad variety of genes and genetic variant types in a single streamlined assay, we reasoned that ONT targeted LRS could be used to improve the diagnosis of spastic-ataxia spectrum disorders. To test this, we designed a targeted assay for evaluation of STR expansions, SNVs, indels, SVs and CNVs across all diagnostically relevant genes (n = 469). We applied this to a cohort of genetically undiagnosed patients with a spastic-ataxia spectrum phenotype and assessed the potential to improve the rate of diagnosis, relative to traditional testing. Here we report the clinical and genetic findings from this cohort, demonstrating a strong improvement in diagnostic rate and highlighting the strengths of LRS for genetic evaluation of these disorders.

## Materials and methods

### Patient recruitment, clinical evaluation and study approval

A cohort of 34 patients with clinically diagnosed spastic-ataxia spectrum disorders, without a genetic diagnosis, were recruited from the Neuromuscular Clinic at Concord Repatriation General Hospital in Sydney, Australia, between April 2023 and March 2024. The cohort consisted of 11 individuals who were naïve to genetic testing and 23 individuals who had previously undertaken genetic testing, as per standard clinical practice in Australia. Prior genetic testing had been arranged by the patients’ treating clinicians, and included various combinations of testing for STR expansions in selected spinocerebellar ataxia (SCA) genes (SCA 1, 2, 3, 6, 7, 12, 17), *FXN* for Friedreich ataxia, *ATN1* for dentatorubral-pallidoluysian atrophy, *FMR1* for Fragile X Tremor/Ataxia Syndrome (FXTAS); NGS (targeted gene panel, WES or WGS); and auxiliary testing (single gene testing, MLPA, microarray, mitochondrial genome sequencing), as appropriate. An additional five individuals with spastic-ataxia spectrum disorders and a known genetic diagnosis were included as positive controls. Demographic, clinical, neuroimaging, neurophysiological and genetic testing data were collected at the time of clinical evaluations and from patient health care records. MRI and/or CT reports, as well as original images, if available, were reviewed. Written informed consent was obtained from all participants according to the Declaration of Helsinki. Study approval for this project was provided by the local Human Research Ethics Committee (2019/ETH12538).

### Targeted long-read sequencing workflow

Peripheral blood samples were obtained from all 39 individuals (34 study participants and five controls). Samples were processed at The Garvan Institute of Medical Research in Sydney, Australia. High–molecular weight (HMW) genomic DNA was extracted from peripheral blood samples using the Nanobind CBB kit (PacBio, Cat# 102-301-900). HMW genomic DNA was sheared to ∼30-kb fragment size using the Diagenode Megaruptor 3 DNA shearing system and visualised on an Agilent Femto Pulse using the Genomic DNA 165 kb Kit. ONT libraries were prepared from ∼3 µg of sheared HMW genomic DNA using a ligation prep (SQK-NBD114.24). Three samples were barcoded and pooled into one library and loaded on an ONT PromethION R10.4.1 flow cell (FLO-PRO114M) and sequenced on either a PromethION 2 Solo or PromethION 48 instrument, with live target selection/rejection executed by the Readfish software package (v0.0.10dev2)^18^ targeting a custom panel of 469 genes associated with spastic-ataxia spectrum disorders, plus the mitochondrial genome (Supplementary Table 1). Libraries were run for 72 hours, with nuclease flushes and library reloading performed at approximately 24 and 48-hour time points.

### Analysis of genetic variation

Raw ONT sequencing data was converted to BLOW5 format^19^ and base-called with Dorado, using the ‘super-accuracy’ model (dna_r10.4.1_e8.2_400bps_5khz_sup_prom.cfg) and the Buttery-eel wrapper to enable BLOW5 input^20^. The resulting ‘pass’ FASTQ files were aligned to both the hg38 and T2T-CHM13v2.0 reference genomes using minimap2 (v2.14-r883)^21^. SNVs and indels were called using Clair3 (v1.0.4)^22^ and phased using WhatsHap (v2.1)^23^. SVs (including CNVs) were called with Sniffles2^24^ (v2.07), which takes haplotagged BAM alignment files to generate phased SV calls. SNVs and indels were functionally annotated using Variant Effect Predictor (VEP)^25^ (v110) and SVs were annotated using AnnotSV^26^ (v3.3.4).

Variants were prioritized for further consideration based on the following criteria: (i) those annotated in ClinVar with a clinical significance of “pathogenic,” “likely pathogenic,” or “uncertain“; (ii) variants predicted to cause a frameshift; (iii) missense variants with a REVEL score greater than 0.5 and classified as “pathogenic”, “likely pathogenic”, “uncertain” or without an assigned clinical significance; and (iv) variants with high or moderate impact that are similarly classified as “pathogenic”, “likely pathogenic”, “uncertain” or lacking an assigned clinical significance in ClinVar. All pre-selected variants were within MANE Select transcripts and either were not found in gnomADv4 or had allele frequency of less than 0.1, resulting in a list of rare and potentially causative variants. Additionally, structural variants intersecting protein-coding genes were also prioritized for further consideration.

### Analysis of STR expansions

We analysed STRs in 21 genes associated with spastic-ataxia spectrum disorders (Supplementary Table 2). During this project, STR expansions in *THAP11* and *ZFHX3*, were identified to cause HCA^27–29^, and these genes were subsequently added to our target panel. All individuals underwent sequencing of *THAP11*, however, 24 samples had already undergone sequencing prior to the addition of *ZFHX3*, and therefore, these individuals were not evaluated for SCA4. Furthermore, *TBP* was omitted from the initial panel, and therefore also was only evaluated in 15/39 samples.

STRs were genotyped using a method demonstrated previously^16^. Each STR region was first inspected in Integrated Genome Viewer (IGV; T2T-CHM13v2 reference)^30^. For sites showing evidence of expansions (i.e. large insertions within alignments spanning an STR site), reads were retrieved within a 50kb window centred on the target STR and assembled *de novo* with Flye (v2.8.1-b1676)^31^ to create a pseudo-haploid contig encompassing the STR region. The starting reads were then realigned to this contig and phased into separate haplotypes using Longshot (v0.4.1)^32^. The initial assembled contig was re-polished with reads from each haplotype using Racon (v1.4.0), generating two distinct haploid contigs encompassing the STR site. The precise position of the STR site was identified by mapping 150-bp unique flanking sequences extracted from T2T-CHM13v2.0 using minimap2 (v2.22). The reads from each haplotype were mapped to their respective polished contigs and re-inspected in IGV to identify potential discrepancies in phasing of reads, which could be manually corrected before re-polishing^30^. Custom sequence bar plots were created in Prism to enable visualisation of STR alleles within and between patients (see Figure 1).

**Figure 1.**
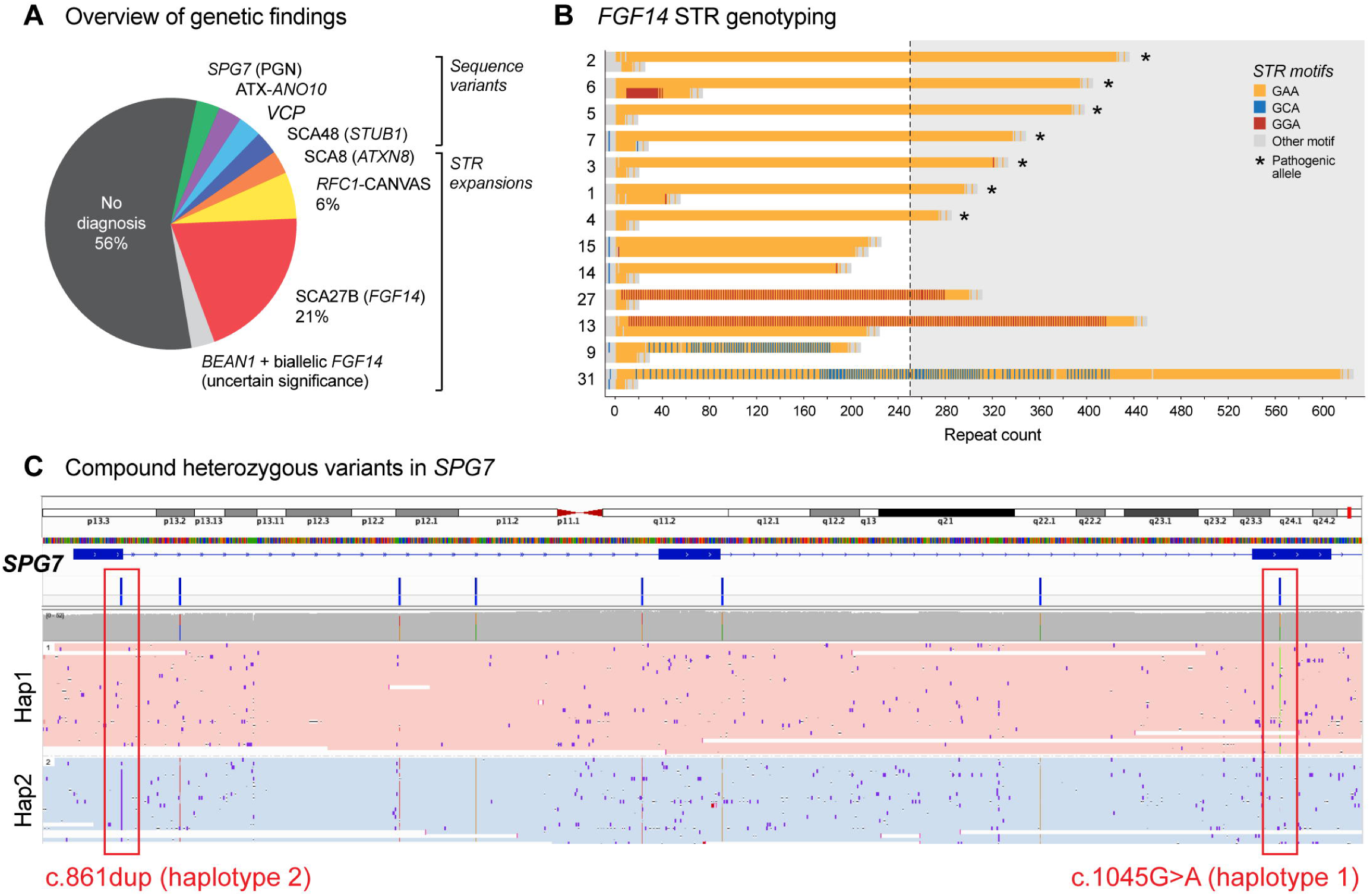
Improved diagnosis of spastic-ataxia spectrum disorders with targeted long-read sequencing. (**A**) Summary of genetic diagnoses in our cohort of 34 genetically undiagnosed patients with spastic-ataxia spectrum disorders. (**B**) Sequence bar charts show STR alleles identified in 13 patients with expanded STRs in *FGF14*. Seven individuals, denoted with asterisks, had pure (GAA) expansions longer than 250 copies, sufficient for SCA27B diagnosis. The remaining six patients had (GAA) expansions that did not reach 250 copies, or expanded (GAA) repeats interrupted by other motifs (GCA or GGA), not considered pathogenic. (**C**) Genome browser view shows detection of two pathogenic heterozygous variants within *SPG7* in a single patient. Alignments are phased into separate haplotypes (pink = haplotype 1; blue = haplotype 2), confirming the two pathogenic variants are on alternative haplotypes (i.e. in trans).

### Validation of results

Our analysis method for genotyping STR expansions with ONT data has previously been validated in a cohort of 37 individuals including patients with neurogenetic diseases, premutation carriers and controls^16^. In the present study, five individuals with a known genetic diagnosis were included as positive controls: one individual with FXTAS due to a *FMR1* STR expansion; one with SCA3 due to a *ATXN3* STR expansion, one with HSP due to homozygous *SPG7* variants; one with recessive ataxia due to compound heterozygous variants in *ANO10;* and one with recessive ataxia due to compound heterozygous variants in *TDP2*. In nine individuals with a potential new genetic diagnosis identified using LRS, findings were confirmed on orthogonal testing completed in laboratories independent to the primary research testing centre. This was undertaken in clinically accredited laboratories for eight individuals, and one individual had completed WGS on a research basis.

### Statistical analysis

Quantitative variables were described as means and ranges. Categorical variables were expressed with frequencies and percentages. Comparison of groups was undertaken using independent samples _X_2 testing for binomial variables. Statistical significance threshold was set at p-value <0.05.

### Data availability

Anonymised data that support the findings of this study are available from the corresponding author, upon reasonable request.

## Results

### Targeted LRS assay for spastic-ataxia spectrum disorders

We developed an ONT ReadUntil targeted LRS assay^18^ designed to capture the full suite of genetic features implicated in spastic-ataxia spectrum disorders in a single assay. This encompasses 469 genes considered diagnostically relevant plus the mitochondrial genome (Supplementary Table 1). Genes included in the targeted panel were selected via detailed literature review^4^ and review of gene panels for ataxia, spastic-ataxia and HSP through PanelApp, and other genetic testing laboratories including Invitae, Blueprint Genetics, PathWest and VCGS^33^. We designed target regions covering each gene and flanking regions 50 kb upstream and downstream to ensure all exons, introns, gene promoters, untranslated regions (UTRs) and other local regulatory sequences are captured. This panel covers STR sites in 21 genes where expanded alleles are known to cause spastic-ataxia spectrum disorders (Supplementary Table 2). In total, the target panel covers 91.8 Mbases or 3.0% of the human genome sequence.

Targeted LRS enables multiple patient samples to be analysed in a single experiment to reduce cost, while still obtaining sufficient coverage depth for accurate and comprehensive analysis. Our study cohort (see below) was sequenced at a rate of three patients per flow cell (ONT PromethION). We obtained median 42-fold coverage depth across target gene regions, with a median absolute deviation (MAD) of 16.79. The coverage depth across individual target regions ranged from a minimum median coverage of 23 to a maximum median coverage of 50 (Supplementary Figure 1A,B).

To validate our approach, positive controls were analysed by blinded investigators (see Materials & Methods). Our targeted ONT LRS approach identified STR expansions in *FMR1* and *ATXN3* in patients with known FXTAS and SCA3, respectively, with relatively similar repeat lengths identified compared to that generated by standard clinical testing (85 vs 87 copies for *FMR1*; 73 vs 76 for *ATXN3*; Supplementary Figure 2A-B). Additionally, ONT LRS was able to identify the presence and position of two AGG interruptions within the *FMR1* CGG STR expansion (Supplementary Figure 2B). The presence of AAG interruptions are known to influence the propensity of the *FMR1* CGG STR expansion to further expand in subsequent generations^34^. We were able to successfully identify SNVs and small indels in 3 positive control patients with recessive spastic ataxic disorders (Supplementary Figure 2A,C). Our ONT LRS approach also allowed for phasing of these variants, confirming that a patient with an apparently homozygous SNV in *SPG7* did have the same variant on two different alleles and confirming that patients with two variants in *ANO10* and *TDP2* carried these variants *in trans* (Supplementary Figure 2C). Interestingly, the patient with known autosomal recessive spinocerebellar ataxia due to variants in *TDP2* was also found to carry a (GAA)_240_ STR expansion in *FGF14*, which is close to the (GAA)_250_ threshold that is currently considered pathogenic.

### Cohort characteristics

We evaluated the performance of our targeted LRS strategy on a cohort of 34 patients with genetically undiagnosed spastic-ataxia spectrum disorders, in addition to five positive controls (see above). In the study cohort, mean age of individuals at the time of review was 64.0 years (range 24-84), with mean age of onset 53.0 years (range one month to 80 years) and mean disease duration of 10.9 years (range 1-37 years). 16/34 (47%) were female. The majority (20/34, 59%) were white/European; 2/34 (6%) Lebanese; 1/34 (3%) each of Chinese, Filippino, Indian and Iraqi; 2/34 (6%) were of mixed backgrounds; whilst ancestry was unknown in 6/34 (18%). A family history of a similar phenotype was present in 12/34 (35%); inheritance was presumed to be autosomal dominant in 11/34 (32%) and autosomal recessive in 1/34 (3%). The predominant clinical phenotypic classification was ataxia in 29/34 (85%), spastic-ataxia in 3/34 (9%) and spastic paraparesis in 2/34 (6%) (Table 1).

**Table 1.**
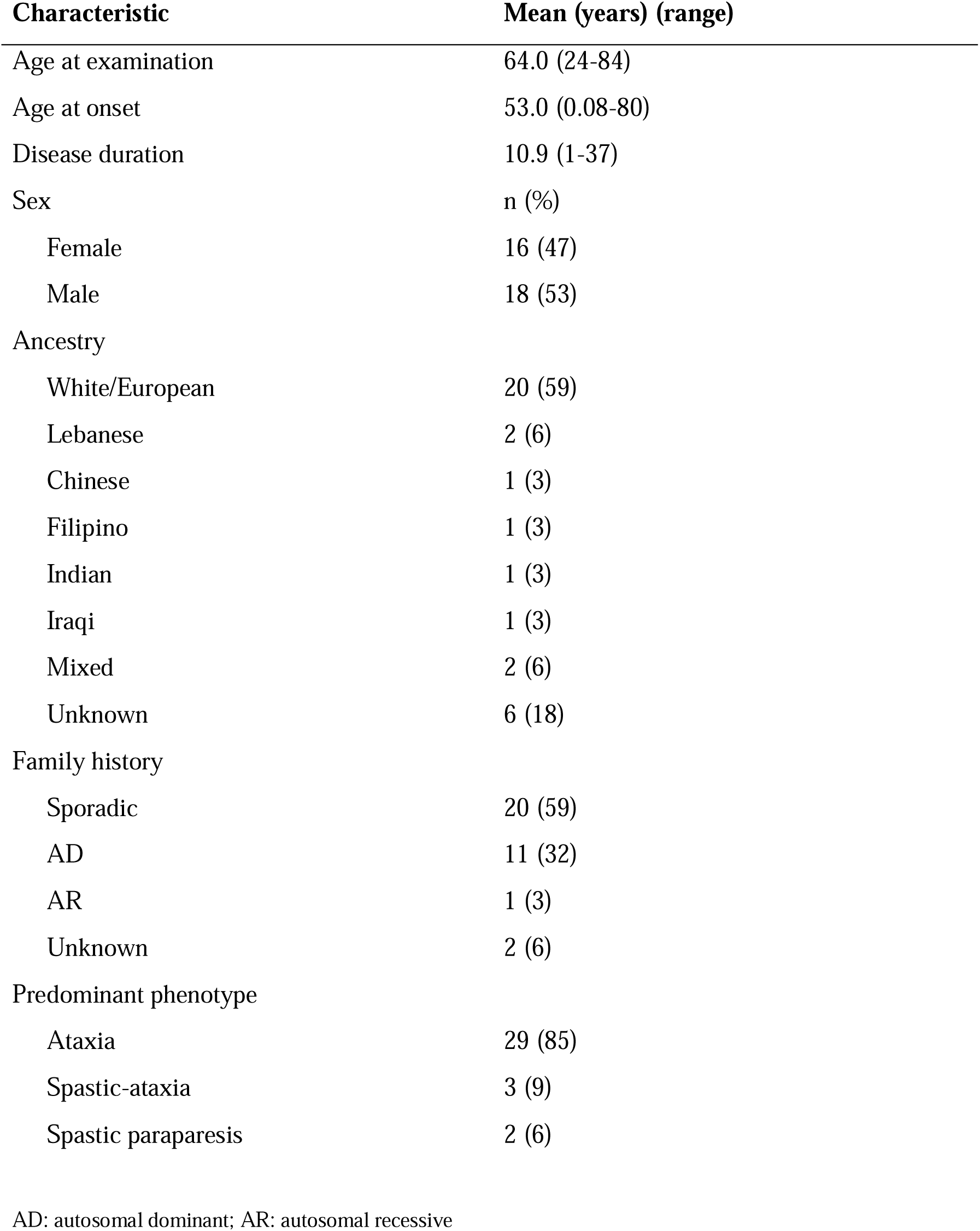
Baseline characteristics of patients with genetically undiagnosed spastic-ataxia spectrum disorders.

| **Characteristic** | **Mean (years) (range)** |
| --- | --- |
| Age at examination | 64.0 (24-84) |
| Age at onset | 53.0 (0.08-80) |
| Disease duration | 10.9 (1-37) |
| Sex | n (%) |
| Female | 16 (47) |
| Male | 18 (53) |
| Ancestry |  |
| White/European | 20 (59) |
| Lebanese | 2 (6) |
| Chinese | 1 (3) |
| Filipino | 1 (3) |
| Indian | 1 (3) |
| Iraqi | 1 (3) |
| Mixed | 2 (6) |
| Unknown | 6 (18) |
| Family history |  |
| Sporadic | 20 (59) |
| AD | 11 (32) |
| AR | 1 (3) |
| Unknown | 2 (6) |
| Predominant phenotype |  |
| Ataxia | 29 (85) |
| Spastic-ataxia | 3 (9) |
| Spastic paraparesis | 2 (6) |

AD: autosomal dominant; AR: autosomal recessive

### Genetic findings

Our targeted LRS strategy identified a causative pathogenic variant sufficient for diagnosis in 14/34 (41%) of participants (Figure 1A). In the group who had previously undergone unsuccessful genetic testing, a diagnosis was obtained in 9/23 (39%) participants, whilst a diagnosis was obtained in 5/11 (45%) who were naive to genetic testing. A heterozygous GAA-*FGF14* STR expansion ≥250 repeats, consistent with a diagnosis of SCA27B, was identified in 7/34 (21%) of the study cohort and was found in 5/23 (22%) of the testing-negative group (Figure 1B). *RFC1* CANVAS/spectrum disorder due to biallelic *RFC1* STR expansions was identified in 2/34 (6%). A single individual was identified to have each of: a CAG·CTG STR expansion in *ATXN8/ATXN8OS*; a homozygous splicing variant in *ANO10* (c.1163-9A>G); a heterozygous missense variant in *VCP* (c.475C>T, p.Arg159Cys); a heterozygous in-frame deletion in *STUB1* (c.433_435del, p.Lys145del); one instance of compound heterozygous missense and nonsense variants in *SPG7* (c.1045G>A, p.Gly349Ser and c.861dup, p.Asn288Ter; Supplementary Figure 3A-D). For the latter case, variants were phased with LRS without parental sequencing data. The 75.7 kb phased block contained six additional heterozygous variants between the two variants of interest, supported by 23 long reads, providing robust evidence to establish compound heterozygosity (Figure 1C). Genetic and clinical features for individuals with a positive diagnosis are summarised in Supplementary Table 3.

This left 20/34 (59%) of individuals without a genetic diagnosis. Two of these individuals were found to harbour pathogenic/likely pathogenic variants in *GCH1* and *PRRT2,* respectively, although these findings were of uncertain clinical relevance to the cerebellar ataxia phenotype. A further individual was identified to have a STR expansion in *BEAN1* in addition to biallelic STR expansions in *FGF14*, within the range of 200-249 repeats. It is likely that one or both of these are the causative genetic events for this patient, however, we currently consider this outcome to be uncertain (see below). Overall, one or more variants of uncertain significance (VUS) were identified in 6/20 (30%) of individuals without an identified causative variant (Supplementary Table 4).

To validate our findings, eight individuals (patients 1, 3, 4, 5, 7, 8, 12 and 13) underwent confirmatory testing through an independent clinically accredited laboratory. One further individual (patient 11) had previously undergone WGS on a research basis and data was available for reanalysis. Five individuals with *FGF14* STR expansions, as well as one individual with biallelic *RFC1* STR expansions underwent confirmatory testing with standard flanking PCR and repeat-primed PCR, followed by capillary electrophoresis. Results demonstrated similar GAA repeat lengths between the two methods and in all cases the pathogenicity of expansions were correctly classified (Supplementary Table 5). The biallelic *RFC1* AAGGG STR expansion identified in patient 8 was also confirmed with standard flanking PCR and repeat-primed PCR followed by capillary electrophoresis, although the specific repeat length was not reported. The homozygous *ANO10* variant identified in patient 12, and the heterozygous *VCP* variant identified in patient 13 were confirmed on clinically accredited WES. The *STUB1* in-frame deletion in patient 11 was confirmed on retrospective review of prior research WGS data, and further confirmatory testing with a clinically accredited WES is pending.

### Clinical findings

#### *FGF14*-GAA (SCA27B); demographic and clinical features

A total of 7/34 (21%) individuals were identified to have GAA≥_250_ *FGF14* STR expansions, consistent with a diagnosis of SCA27B. Mean GAA repeat length was 348 repeats (range 274-425, SD +/−56) (Figure 1B). All seven individuals were male. Ancestry was known in 6/7 (86%) individuals, with white/European ancestry in all known cases. An autosomal dominant family history was apparent in 3/7 (43%) of individuals, with maternal inheritance in all cases. Mean age at examination was 75.4 years (range 69-81), mean age of onset was 65.3 years (range 53-75) and mean disease duration was 10.1 years (range 6-20). 4/7 (57%) required a mobility aid (stick 2/4 and walker 2/4), with a mean time to mobility aid requirement of 5.8 years from disease onset (range 4-10). Mean SARA score was 11.6 (range 6.5-18). Clinical features of all genetically diagnosed patients, and those with SCA27B specifically, are indicated in Figure 2A,B.

**Figure 2.**
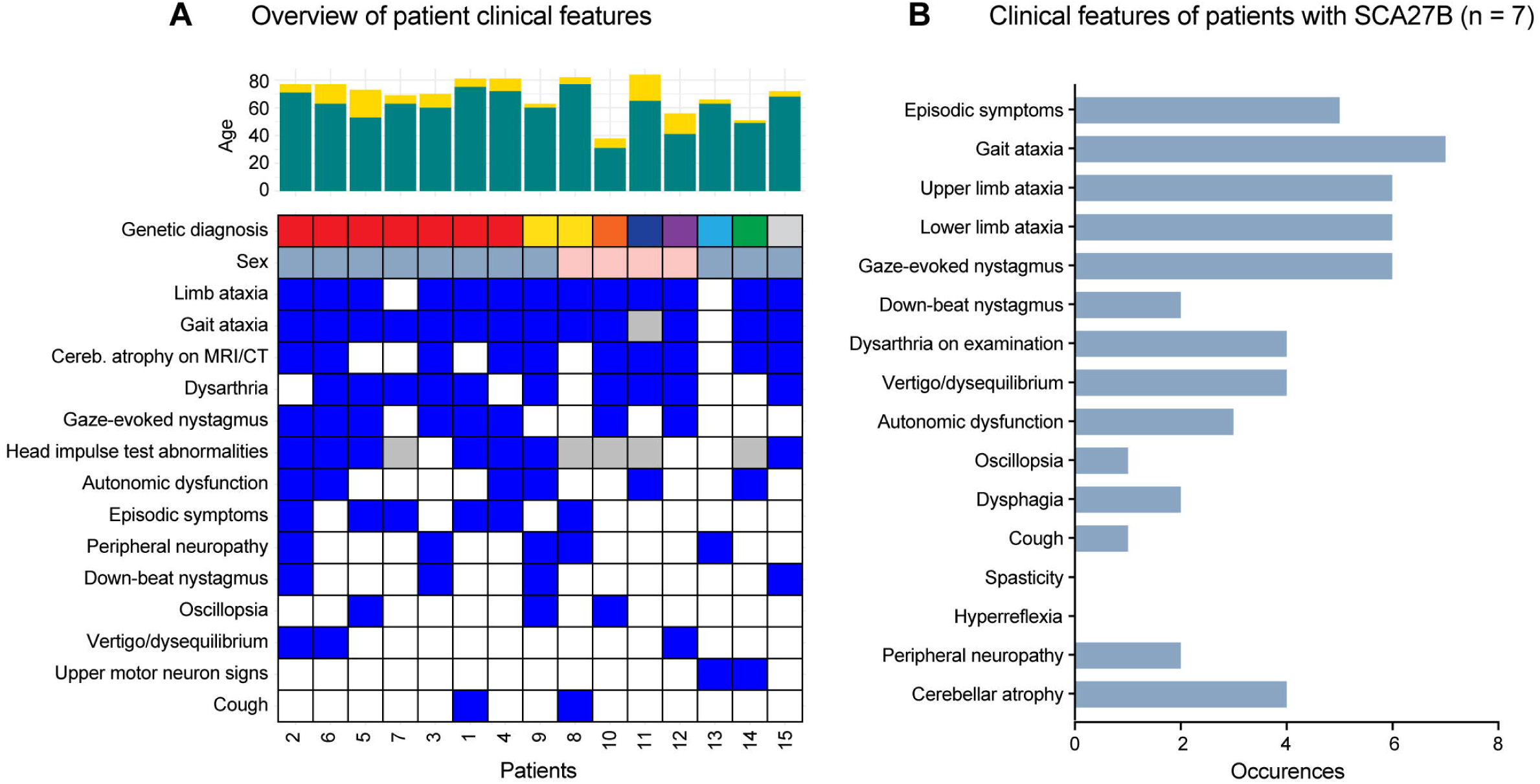
Clinical features of spastic-ataxia patients receiving a genetic diagnosis via long-read sequencing. (**A**) Chart provides an overview of clinical characteristics for all patients who received a genetic diagnosis (plus patient#15, for whom two possible pathogenic STR expansions were detected). The upper bar chart shows patient ages at time of analysis (yellow) and their age at symptom onset (green). The heatmap below indicates symptoms identified in each patient (blue = positive; white = negative; grey = not assessed). Patient sex is encoded with blue (male) or pink (female) tiles and genetic diagnoses are coded as follows: *FGF14* = red; *RFC1* = yellow; *ATXN8* = orange; *STUB1* = navy; *VCP* = blue; *ANO10* = purple; *SPG7* = green; *BEAN1* and biallelic *FGF14* (uncertain clinical significance) = grey. Patients with SCA27B are sorted by *FGF14* expansion size, in matched order to Figure 1B. (**B**) Bar chart summarises the frequency of clinical phenotypes observed among patients diagnosed with SCA27B, based on STR expansions in *FGF14* (n = 7 patients).

#### Non-pathogenic *FGF14* STR expansions

Four individuals were identified to have non-canonical interrupted GAA *FGF14* STR expansions, three of which were ≥250 copies in length (Figure 1B). The longest was a mixed GAA/GCA expansion of 616 repeats, with a maximal uninterrupted GAA length of 195 repeats, while two patients had mixed GAA/GGA repeats with total repeat lengths of 440 and 330, respectively. None of these non-canonical alleles contained an uninterrupted stretch of ≥250 GAA copies and, as a result, none were considered to be pathogenic.

Two further participants, as well as one of the positive controls, were found to have GAA repeats in the 200-249 range (Figure 1B). Patient 13 had a STR expansion of 213 pure GAA repeats, in addition to the non-pathogenic mixed 440 GAA/GGA repeat. This individual was identified to have a pathogenic variant in *VCP*, with a matching phenotype, and the *FGF14* GAA_213_ STR expansion was not felt to be contributory to his presentation. The other individual was identified to have both a biallelic *FGF14* GAA repeat expansion, with 213/201 repeats, as well as a STR expansion in *BEAN1*, both of which may contribute to their phenotype. This case is discussed in detail below.

#### *RFC1*-CANVAS; clinical and genetic features

*RFC1*-CANVAS was identified in two individuals. Both presented with the full CANVAS syndrome including cerebellar ataxia, neuropathy and vestibular areflexia, with additional clinical features indicated in Supplementary Table 3. Patient 8 was of white/European background and had biallelic (AAGGG)_n_ expansions, with 651 and 693 repeats. The second individual (patient 9), from the Philippines, was compound heterozygous for an (AAGGG)_1000_ expansion and an (ACAGG)_2000_ expansion.

#### Non-pathogenic *RFC1* STR expansions

Two individuals had monoallelic *RFC1* (AAGGG)_n_ expansions of 483 and 440 repeats respectively (Figure 2A). Prior reports have identified that compound heterozygosity with a *RFC1* (AAGGG)_n_ expansion and a truncating variant can cause *RFC1*-CANVAS^35,36^. However, ONT LRS sequencing data did not identify a second hit in either of these two cases.

#### Two suspicious STR expansions (*FGF14/BEAN1*) in the same individual

A man of European background presented with sporadic late-onset ataxia from age 66-70 years. The year prior to symptom onset, the patient had suffered a stroke with residual speech and swallow impairment. He had a single episode of vertigo at age 66-70, attributed to benign paroxysmal positional vertigo, but episodic features were not otherwise reported. Two years post symptom onset, he required use of a walker, and after a further year, commenced using a wheelchair. On examination, he had down-beat nystagmus, saccadic pursuit, dysmetric saccades, dysarthria, upper limb dysmetria, dysdiadochokinesia, heel-shin ataxia and an ataxic gait. Head impulse test was positive to the left. MRI demonstrated generalized cerebral and cerebellar atrophy, in addition to mineral deposition in the basal ganglia and a chronic infarct in the posterior limb of the left internal capsule. ONT LRS identified a STR expansion in *BEAN1* consisting of (TAAAA)_50_(TGGAA)_226_(TAAAA)_120_(TGGAA)_110_(TAAAA)_75_, as well as a biallelic *FGF14* STR expansion with 213/201 GAA repeats.

The pathogenic insertion in *BEAN1* consists of complex penta-nucleotide repeats, including (TGGAA)_n_, (TAGAA)_n_ and (TAAAA)_n_, followed by (TAAAATAGAA)_n_, with the (TGGAA)_n_ component thought to be causative for disease^37^. It is one of the most common causes of autosomal dominant SCA in Japan, but has only been rarely reported elsewhere, including in individuals from China, Taiwan, Korea, and in a Brazilian family with Japanese ancestry^38^. To our knowledge, the pathogenic *BEAN1* expansion has not been previously reported as causing ataxia in an individual of European ethnic background, therefore the clinical relevance of this finding was considered uncertain. Additionally, the *FGF14*-GAA 200-249 alleles were significantly enriched in patients with downbeat nystagmus compared to controls, with a similar phenotype compared to patients carrying a (GAA)≥250 expansion^39^. However, the relevance of the 200-249 expansion is still considered uncertain and requires large-scale studies to confirm the clinical significance^39^. Finally, there is a suggestion that biallelic GAA alleles <250 repeats might be associated with disease^40^ but will also require publication of further cases. Overall, it was uncertain whether the genetic findings in *BEAN1* and *FGF14* were causative or contributing to the clinical phenotype in this individual.

#### Clinically-relevant sequencing variants

We identified clinically-relevant sequencing variants in the *VCP* and *STUB1* genes in unsolved participants (**Figure 1A, 2A**).

The *VCP* variant carrier (patient 13), a man in his 60s of white/European ancestry, presented with features of complex hereditary spastic paraplegia (Supplementary Table 3). He had a history of osteoporosis, migraines and had previously suffered a right cerebellar stroke, with complete symptomatic recovery. There were no cognitive symptoms. Family history revealed that one of his parents had mobility issues and dementia, and his sibling had frontotemporal dementia. MRI demonstrated a small right chronic cerebellar infarct. Cortical excitability studies using threshold tracking transcranial magnetic stimulation (TMS) were performed, demonstrating normal short interval intracortical inhibition (SICI). Abnormal SICI is a biomarker of inhibitory GABAergic circuit dysfunction^41^. There are no publications on the use of TMS in VCP-containing protein disease, however reduced SICI, implying cortical hyper-excitability, is reported in other TAR DNA-binding protein 43 (TDP-43) proteinopathies, such as ALS and FTLD. A dual-phase whole-body bone scan did not demonstrate scintigraphic evidence of Paget’s disease. ONT LRS identified a pathogenic missense variant in *VCP* (c.475C>T, p.Arg159Cys). This was confirmed on clinically accredited WES and has been previously reported^42^.

Patient 11, the *STUB1* variant carrier, a lady in her 80s of European ancestry presented with autosomal dominant ataxia, with similar features reported in one of her parents and mild features in her child. Clinical features are indicated in Supplementary Table 3. ONT LRS identified an in-frame deletion in *STUB1* (c.433_435del). Prior SCA17 testing had identified normal *TBP* repeat length (35/36 repeats). The patient had previously undergone WGS, and data was retrospectively reviewed, with confirmation of the *STUB1* variant. Since the time WGS was originally undertaken, additional cases with the same *STUB1* variant have been published, permitting classification of the variant as likely pathogenic^5,43^.

#### Sequencing variants of uncertain clinical relevance

Variants in the *GCH1* and *PRRT2* genes were identified in unsolved participants (Supplementary Table 4), despite follow-up examinations the clinical relevance was unclear.

The *GCH1* variant carrier was a lady in her 60s, of mixed ancestry, who presented with progressive balance decline, dizziness and an upper limb tremor from age 61-65. There was no family history of similar features. Dizziness was constant, with exacerbations triggered by crowds or emotion, lasting 5-40 minutes, as well as brief instantaneous exacerbations with turning the head. On examination there were mild features of ataxia, with saccadic pursuit, horizontal gaze-evoked nystagmus, intention tremor, mild heel-shin ataxia and a wide-based gait. Head-impulse testing was negative. Sensory examination demonstrated mild distal sensory impairment in the lower limbs. There was a jerky tremor of the upper limbs, most notably with certain postures, suspected to reflect a dystonic tremor. There were no other extrapyramidal features. ONT LRS identified a variant in *GCH1* (c.671A>G, p.Lys224Arg), classified as likely pathogenic. Additional VUS were identified in *KCNA1* (c.368G>A, p.Gly123Asp) and *TTBK2* (c.3485G>A, p.Arg1162His).

The *PRRT2* variant carrier, a man in his 50s, with European ancestry, presented with a complex neurologic disorder of ten year duration. He had a family history of dementia in one parent and in one of their parents. Features included fasciculations, spasticity, hyperreflexia and mild distal weakness of upper and lower limbs, cerebellar features (gaze-evoked nystagmus, saccadic pursuit, dysmetric saccades, dysarthria and upper and lower limb ataxia), upper limb bradykinesia and a mild rest tremor. Gait was broad-based, with slow initiation and shuffling steps. He reported autonomic features (erectile dysfunction and urinary urgency) and although not formally tested, cognitive decline was suspected, with difficulties processing complex information, and a tendency to burst into laughter with cognitive loading, suggestive of pseudobulbar affect. MRI demonstrated diffuse cerebral involution without disproportionate cerebellar atrophy. NCS revealed a sensorimotor polyneuropathy, and EMG showed chronic neurogenic changes in limb and cranial nerve-innervated muscles. Motor evoked potentials revealed marked dysfunction of central motor pathways to all limbs. During investigation, the patient was also identified to have positive CASPR2 antibodies on blood testing, which were negative on CSF. This finding was of uncertain significance, but the patient was trialled on intravenous immunoglobulin and mycophenolate without significant response. ONT LRS identified a variant in *PRRT2* (c.649dup, p.Arg217ProfsTer8), which was classified as pathogenic although is of uncertain clinical significance in this case. An additional VUS was identified in *SPTBN2* (c.6250G>A, p.Glu2084Lys).

## Discussion

Our study evaluated 34 patients with genetically undiagnosed spastic-ataxia spectrum disorders using targeted ONT LRS and identified a likely diagnosis in 14/34 (41%; summarised in Figure 3). In the group who had not previously undergone genetic testing, the diagnostic yield was 45% (5/11). In those who had previously had genetic evaluation, with no diagnosis found, ONT LRS identified a cause in 39% (9/23). The high success rate in the latter group demonstrates the potential for LRS to greatly improve the rate of genetic diagnosis in these disorders.

**Figure 3.**
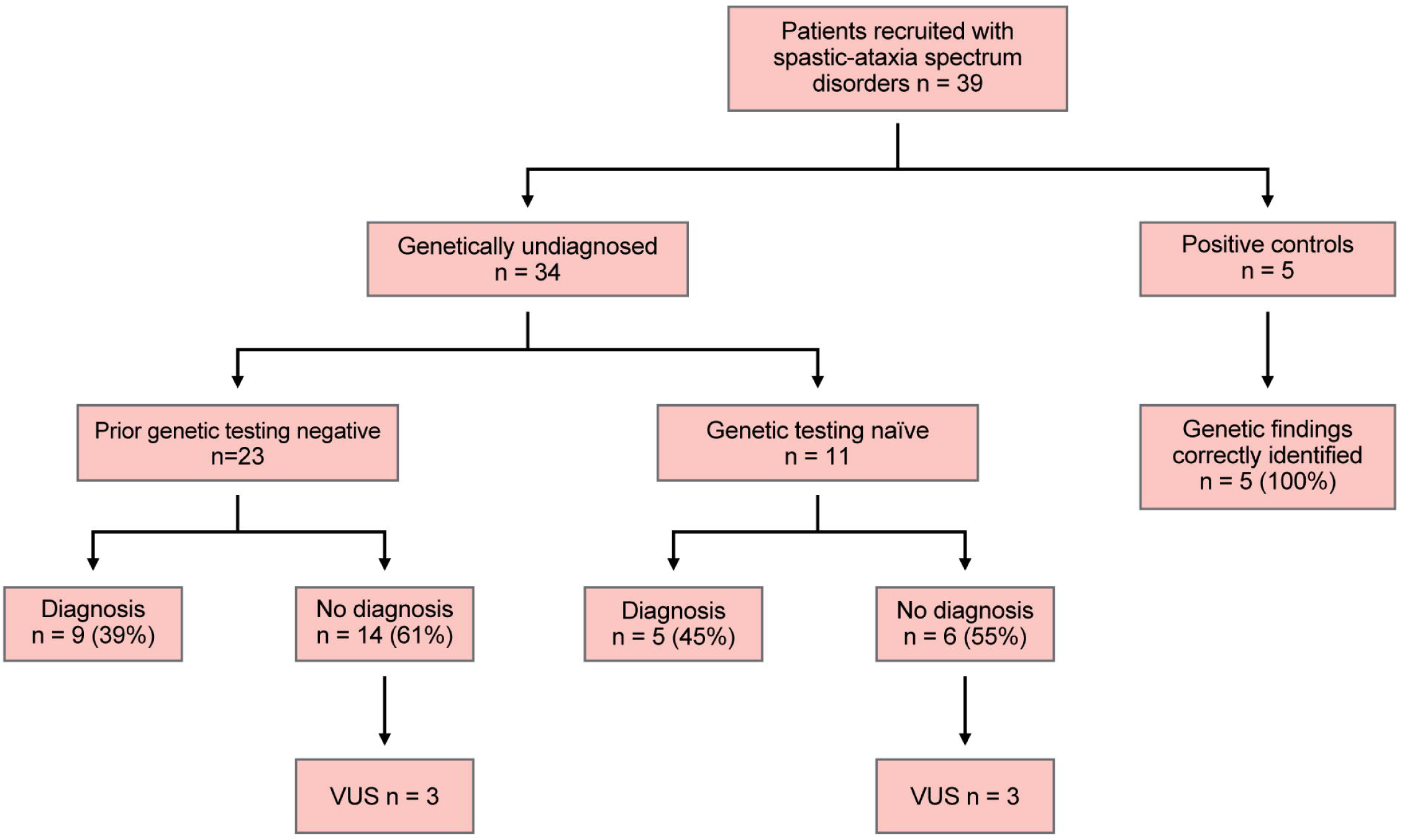
Overview of outcomes. Flow chart provides a schematic overview of diagnostic outcomes across the study cohort, which included 34 genetically undiagnosed patients with spastic-ataxia spectrum disorders and five patients with known genetic diagnoses included as positive controls.

There are multiple explanations for the diagnostic uplift achieved by our LRS assay. STR expansions account for most diagnoses in the cohort (10/14, 71%). The most common diagnosis was SCA27B, identified in 7/34 (21%), whilst two individuals (6%) were identified to have *RFC1*-CANVAS. These two STR expansions were both only relatively recently identified^44–46^, and currently, testing in Australia is only offered on a research basis. Lack of access to clinical testing combined with the relatively high frequency of *FGF14* and *RFC1* STR expansions in patients with HCA underpins the high rate of detection of these two variants within our cohort. Another individual was diagnosed with SCA8, caused by an STR expansion in *ATXN8*/*ATXN8OS*^47^. While clinical testing is available, this is one of several rare STR expansions that are not incorporated into standard genetic testing pathways in all sites, so may be overlooked, as was the case for this individual. The ability to identify these unexpected diagnoses is an advantage of our integrated approach that captures all relevant STRs on every test. The remaining 4/14 (29%) of diagnoses were accounted for by conventional sequence variants (SNVs and indels). Notably, this includes a compound heterozygous variant in *SPG7*, which could not be phased using short-read NGS to confirm pathogenicity. The capacity of LRS to phase autosomal recessive or *de novo* variants or identify both an STR expansion and sequence variant in a compound heterozygous state (as can occur in *RFC1-*CANVAS/spectrum disorder and Friedreich ataxia^35,48^), is especially valuable for late-onset conditions, where availability of both parents for testing may otherwise be a barrier to diagnosis. Finally, one patient was diagnosed with SCA48 caused by a variant in *STUB1*, which was not recognised as pathogenic at the time of previous genetic evaluation due to the lack of recognition of dominant inheritance of this gene at the time of initial analysis^49^.

Pathogenicity of STR expansions may depend not only on repeat length, but on the repeat motif composition and the presence of interruptions. For example, *RFC1-*CANVAS/spectrum disorders are usually caused by biallelic (AAGGG)_n_ STR expansions, but there are other pathogenic motifs, including (ACAGG)_n_, as well as several non-pathogenic motifs, including (AAAAG)_n_, (AAGAG)_n_, (AAAGGG)_n_ and smaller (AAAGG)_n_ expansions^5^. In such disorders, it is vital to obtain both repeat length and sequencing data, in order to confirm a diagnosis. Clinical testing may be undertaken with a combination of flanking PCR and repeat-primed PCR, which will identify specific common repeat motif(s), but will typically not detect the less common motifs, requiring further testing for such cases^50,51^. In contrast, LRS is able to detect and characterise the various repeat motif types, as well as repeat length in a single test. For example, patient 9, with *RFC1-*CANVAS, was identified to have both an (AAGGG)_n_ and an (ACAGG)_n_ expansion. Similarly, in another patient, we identified a *BEAN1* STR expansion and were able to resolve the complex motif configuration; (TAAAA)_50_(TGGAA)_226_(TAAAA)_120_(TGGAA)_110_(TAAAA)_75_. Notably, this also reflects the first known case of European ancestry to harbour the pathogenic (TGGAA)_n_ motif. Furthermore, several individuals were found to have non-pure GAA *FGF14* STR expansions. In these cases, LRS provided the capacity to quantify the maximum pure GAA repeat length, which fell below the 250-repeat threshold for pathogenicity in all cases^40^. This highlights the strong utility of LRS for evaluation of STR expansions, being the only technique that can identify size, motif composition, zygosity, presence of interruptions, flanking variation and methylation status in a single assay^16^.

The ability to capture the full variety of genomic features potentially implicated in spastic-ataxia spectrum disorders in a single assay not only improves diagnostic yields but can greatly streamline the diagnostic pathway. In clinical practice, STR expansions are commonly evaluated with repeat-primed PCR and/or Southern blot, which require separate assays and/or specific primers/probes for each different gene^16^. In addition to requiring multiple sequential tests for some patients, this also incurs delays in the availability of clinical testing for newly described STR expansions as each new gene-specific assay must be developed and validated^5^. In contrast, new genes and/or STR targets can be added to an ONT adaptive sampling assay simply by updating the set of genome targets provided programmatically during sequencing, requiring no change to either the upstream laboratory processes or downstream bioinformatics analysis pipelines^18^. Therefore, ONT LRS facilitates adaptability and rapid addition of new genes as they are discovered. This was exemplified in our study, as during the project, two additional ataxia-associated STR expansions were identified; *THAP11* (‘SCA51’)^27,52^ and *ZFHX3* (recently identified as the basis of SCA4)^28,29^. Both genes were subsequently added to our gene panel. However, 24 samples had already undergone sequencing and were not evaluated for STR expansions in *ZFHX3.* Conversely, this also brings to light a limitation of targeted ONT LRS, which is the inability to reanalyse data for variants at additional ‘off-target’ loci, if new genes are discovered. This would be negated using whole-genome LRS, with either ONT or Pacific Biosciences (PacBio) technology but would come at an additional cost per patient.

Despite the described benefits of LRS, there remain several limitations and obstacles to clinical uptake of this technology. On a read-by-read level, accuracy for detecting single nucleotide variants (SNVs) and small insertions/deletions (indels) is lower than NGS^53^. Targeted ONT LRS can identify 98.8% of SNVs, although greater inaccuracy is seen in samples with reduced depth of coverage^53^. Detection of indels suffers from a greater level of inaccuracy, on account of an increased propensity for slippage errors.^18^ Software tools are less well-developed for data analysis, mapping and variant calling compared to NGS^13^. Due to the need for HMW DNA, freshly collected samples and specialized DNA extraction procedures are generally required^13^. Finally, the cost of LRS remains higher than short-read NGS, although pricing has, and is predicted to continue to improve^13^.

Beyond technology-related factors, other limitations of this study include potential selection bias. Patients were recruited from a tertiary centre, receiving referrals predominantly from primary care physicians and neurologists, with recruitment undertaken by authors L.I.R, D.Y. and K.R.K., neurologists, with subspecialty experience in neurogenetic disorders. The application of study protocols to broader neurology clinics could potentially impact the cohort characteristics and diagnostic rate. Most individuals in our cohort were of white/European background, and therefore results may differ when applied to broader population groups. Furthermore, although patients with features consistent with any spastic-ataxia spectrum disorder were eligible for the study, 29/34 (85%) had a primary HCA phenotype, whilst only two individuals had a primary HSP phenotype. This was likely contributed to by referral patterns, with several HCA patients referred to our research centre by local neurologists aware of this study, as well as the commencement of recruitment in late 2023 for a concurrent study, for which individuals with a primary HSP phenotype were eligible.

After undergoing genetic evaluation, many individuals with spastic-ataxia spectrum disorders remain without a genetic diagnosis, which has implications for clinical care. Provision of a genetic diagnosis may terminate a prolonged and arduous diagnostic odyssey, avoid further unnecessary and invasive investigations, permit disease-specific management and provide more accurate prognostic information. Although only few gene-specific treatments are presently available (e.g. treatment with 4-aminopyridine for SCA27B)^39^, eligibility for clinical trials may also be affected by a genetic diagnosis. Furthermore, there are important implications for reproductive choices and family counselling, with the ability to offer genetic testing in at-risk family members^54^. Evidently, there is clear impetus to improve current genetic testing methods, to close this diagnostic gap. Our study begins to address this gap through the development and evaluation of a targeted LRS strategy that accurately identifies all classes of genomic variation across a panel of genes implicated in spastic-ataxia spectrum disorders. This integrated ‘single test’ approach is now poised to replace fragmented genetic testing regimes, leading to improved diagnostic rates and streamlining the diagnostic pathway for patients.

## Data Availability

All data produced in the present study are available upon reasonable request to the authors

## Acknowledgements

We thank our colleague David Pellerin for aiding in interpreting potential atypical cases of SCA27B. Kishore Raj Kumar receives funding from the Medical Research Future Fund (grants 2023126, 2023357, and 2024888) and from the Ainsworth 4 Foundation, unrelated to the current project.

## Supplementary material

Supplementary material is available at online.

