## Supplementary Figure 1 for "Targeted long-read sequencing as a single assay improves diagnosis of spastic-ataxia disorders"

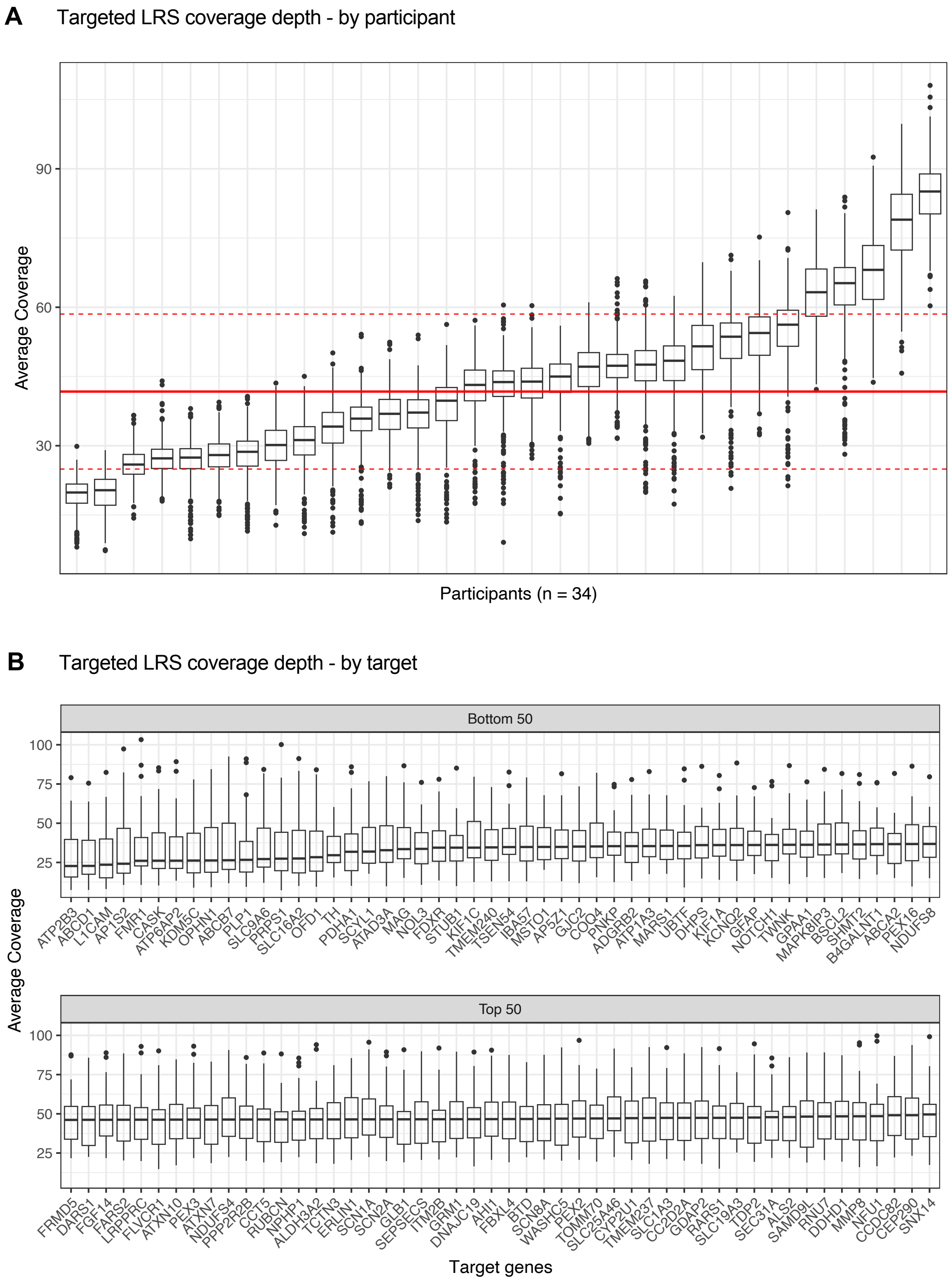


**Supplementary Figure 1. Sequencing coverage metrics for targeted long-read sequencing assay.**
(**A**) Average coverage depth for gene targets (n = 469) obtained for each participant in our undiagnosed spastic-ataxia cohort (n = 34 individuals). (**B**) Average coverage depth for participants (n = 34) obtained for each gene target on our targeted long-read sequencing panel; given space constraints, only the fifty genes with lowest and highest coverage are shown.
