## Supplementary Figure 2 for "Targeted long-read sequencing as a single assay improves diagnosis of spastic-ataxia disorders"

**A** Summary of known pathogenic variants in control cases

| Participant | Diagnosis | Gene | Variant(s) | Original testing method | ONT LRS Result |
|---|---|---|---|---|---|
| 35 | FXTAS | *FMR1* | $(CCG)_{85}$ | PCR and triplet-primed PCR | $(CGG)_9(AGG)_1$ $(CGG)_9(AGG)_1(CGG)_{67}$ |
| 39 | SCA3 | *ATXN3* | $(CAG)_{73}$ | PCR and fragment analysis | $(CAG)_2(CAA)_1(AAG)_1(CAG)_1(CAA)_1(CAG)_{71}$ |
| 36 | SPG7 | *SPG7* | c.1529C>T (p.Ala510Val) presumed homozygous | Targeted NGS panel | c.1529C>T in homozygosity |
| 37 | SCAR10 | *ANO10* | c.132dup (p.Asp45Argfs*9); | Targeted NGS panel | c.132dup (p.Asp45Argfs*9) ; c.1219-1G>T *in trans* |
| 38 | SCAR23 | *TDP2* | c.425+1G>A; c.728del (p.Met243Argfs*3) | WGS | c.425+1G>A ; c.728del (p.Met243Argfs*3) *in trans* |

**B** Identification of STR expansions in control cases

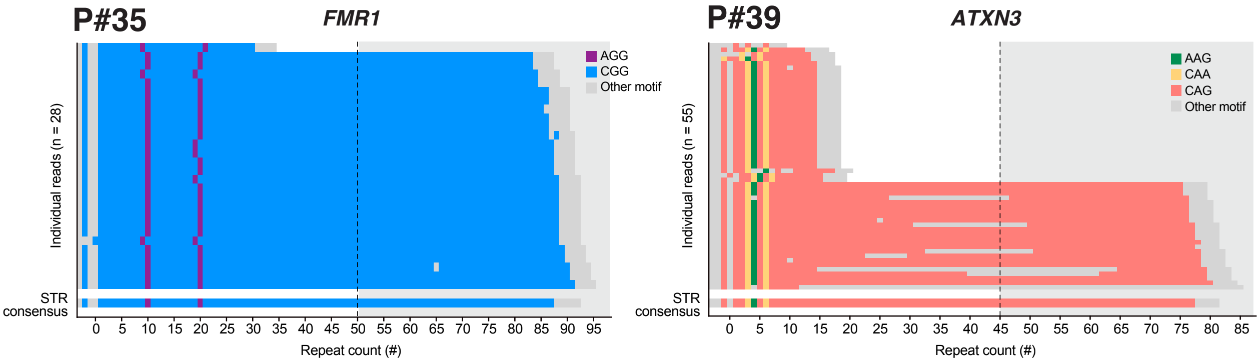


**C** Detection and phasing of sequence variants in control cases

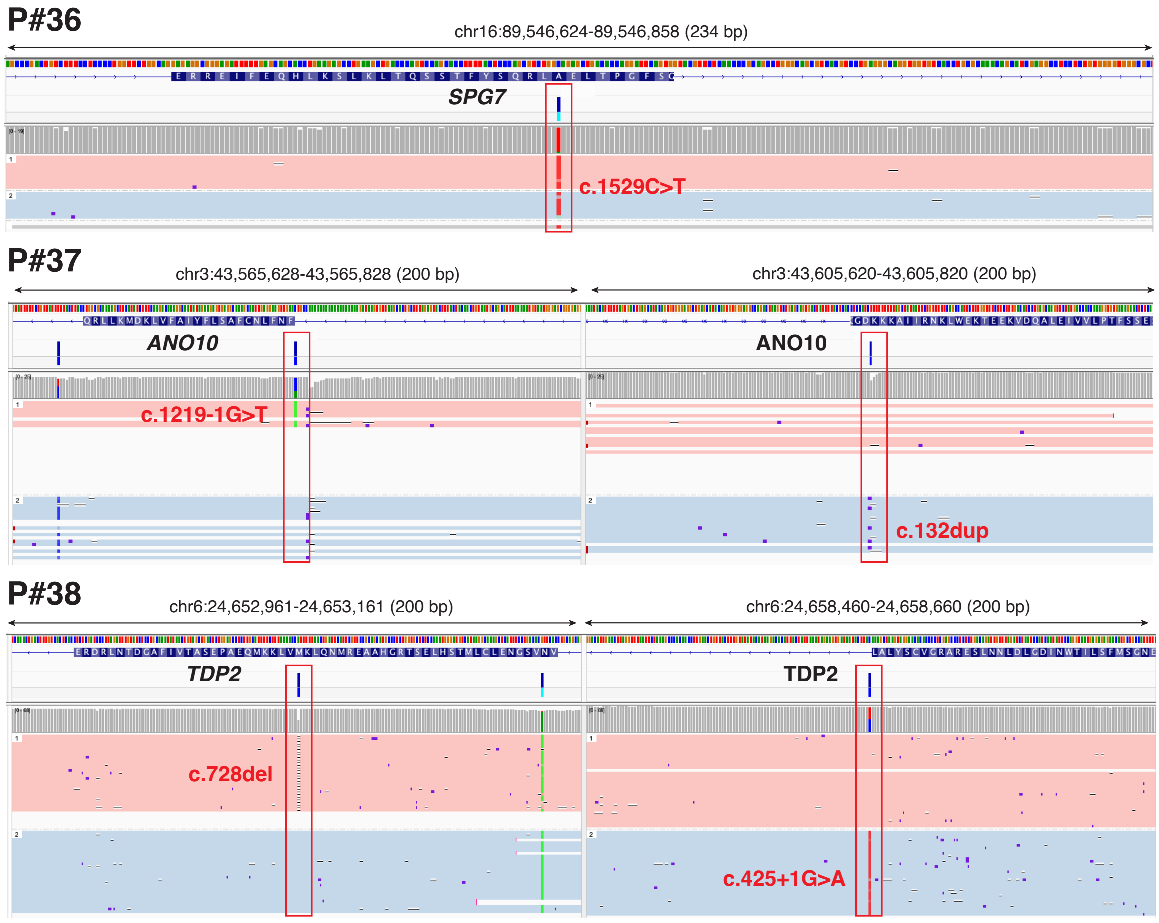


**Supplementary Figure 2. Accuracy of targeted long-read sequencing confirmed by analysis of positive control cases.** (**A**) Table provides an overview of analysis results for five spastic-ataxia patients with known genetic diagnoses who were included as positive controls. Glossary: FXTAS, fragile X tremor/ataxia syndrome; NGS, next generation sequencing; ONT LRS, Oxford Nanopore Technologies long-read sequencing; PCR, polymerase chain reaction; SCA, spinocerebellar ataxia; SCAR, spinocerebellar ataxia, recessive; SPG, spastic paraplegia; WGS, whole genome sequencing. (**B**) Sequence-bar plots show STR genotyping results for FMR1 (Patient #35; FXTAS) and ATXN3 (Patient #39; SCA3). Each individual long-read alignment is show separately and a consensus sequence for the expanded allele in each case is shown below (note: only one allele is present for Patient #35, as FMR1 is on chrX and the individual is male). (**C**) Genome browser view shows detection of known pathogenic sequence variants within SPG7 (Patient #36), ANO10 (Patient #37) and TDP2 (Patient #38). Alignments are phased into separate haplotypes (pink = haplotype 1; blue = haplotype 2), confirming the pathogenic variants for #37 and #38 are on alternative haplotypes (i.e. in trans).
