## Supplementary Figure 3 for "Targeted long-read sequencing as a single assay improves diagnosis of spastic-ataxia disorders"

Detection and phasing of pathogenic sequence variants in four undiagnosed participants

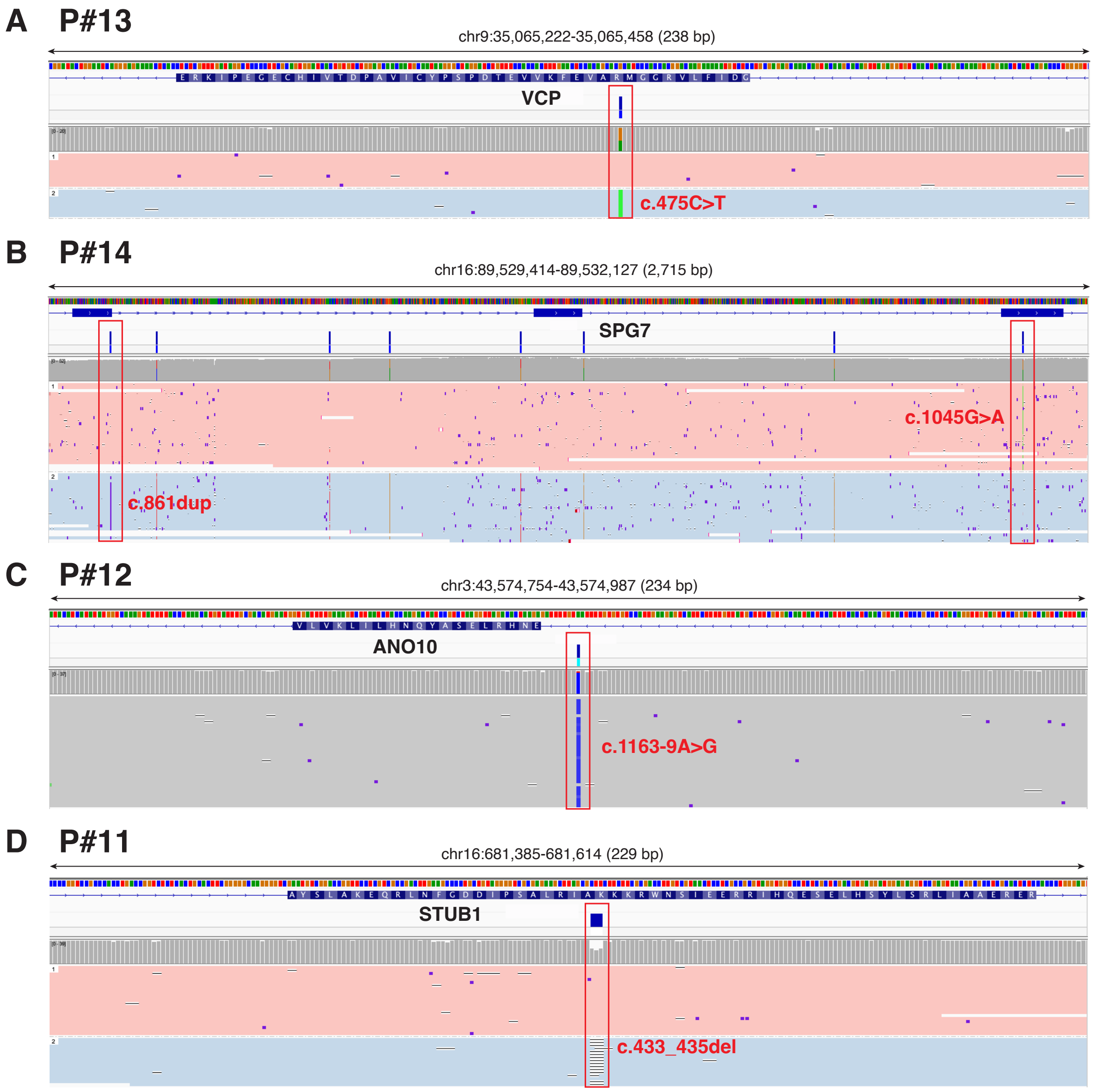


**Supplementary Figure 3. Detection and phasing of pathogenic sequence variants in four undiagnosed participants.** Genome browser view shows detection of pathogenic sequence variants within (**A**) VCP (Patient #13); (**B**) SPG7 (Patient #14); (**C**) ANO10 (Patient #12) and (**D**) STUB1 (Patient #11). Alignments are phased into separate haplotypes (pink = haplotype 1; blue = haplotype 2).
