## Supplementary Table 3 for "Targeted long-read sequencing as a single assay improves diagnosis of spastic-ataxia disorders"

**Table 2. Clinical features and genetic findings for patients with a causative variant**

| **ID** | **1** | **2** | **3** | **4** | **5** | **6** | **7** |
| --- | --- | --- | --- | --- | --- | --- | --- |
| **Diagnosis** | SCA27B | SCA27B | SCA27B | SCA27B | SCA27B | SCA27B | SCA27B |
| **Variant** | *FGF14* GAA expansion (296/40 repeats) | *FGF14* GAA expansion (425/14 repeats) | *FGF14* GAA expansion (321/9 repeats) | *FGF14* GAA expansion (274/9 repeats) | *FGF14* GAA expansion (387/9 repeats) | *FGF14* GAA expansion (394/64 repeats) | *FGF14* GAA expansion (338/17 repeats) |
| **Age (yrs)** | 81-85 | 76-80 | 66-70 | 81-85 | 71-75 | 76-80 | 66-70 |
| **AAO (yrs)** | 71-75 | 71-75 | 56-60 | 71-75 | 51-55 | 61-65 | 61-65 |
| **Disease duration (yrs)** | 6 | 6 | 10 | 9 | 20 | 14 | 6 |
| **Sex** | M | M | M | M | M | M | M |
| **FHx** | Sporadic | Sporadic | AD | Sporadic | AD | AD | Sporadic |
| **Primary phenotype** | Ataxia | Ataxia | Ataxia | Ataxia | Ataxia | Ataxia | Ataxia |
| **Mobility aid** | Stick | Nil | Walker | Nil | Walker | Nil | Stick |
| **1st symptom** | Episodic diplopia | Unsteadiness | Impaired upper limb coordination | Episodic balance impairment | Fall | Unsteadiness, balance impairment | Mobility and balance impairment |
| **Episodic symptoms** | + | + | - | + | + | - | + |
| **Gait ataxia** | + | + | + | + | + | + | + |
| **Limb ataxia** | + | + | + | + | + | + | - |
| **Dysarthria** | + | - | + | - | + | + | +* |
| **Nystagmus** | GE | GE, DBN, UBN | GE, DBN | GE | GE | GE | - |
| **Head impulse** | + | + | - | + (unilateral) | + | + | N/A |
| **Lower limb spasticity** | - | - | - | - | - | - | - |
| **Reflexes** | Normal | Normal | Reduced at ankles | Normal | Normal deep tendon reflexes, upgoing plantar responses | Normal | Normal |
| **Sensory deficit** | - | + | + | - | - | - | - |
| **Autonomic dysfunction** | - | + | - | + | - | + | - |
| **Cough** | + | - | - | - | - | - | - |
| **SARA** | 6.5 | 10.5 | 15 | 10 | 18 | 9.5 | N/A |
| **MRI/CT abnormalities** | - | Mild cerebellar atrophy (vermis-predominant), small right acoustic neuroma | Cerebellar and brainstem atrophy | Cerebellar atrophy (vermis-predominant), mild cerebral atrophy | - | Cerebellar atrophy (most marked in superior vermis), cerebral atrophy | - |
| **NCS/EMG** | Bilateral CTS, otherwise normal | Sensory axonal neuropathy | Sensorimotor polyneuropathy | N/A | N/A | N/A | Normal |
| **Other** |  | Vertigo/dysequilibrium, nocturnal hiccups |  | Postural upper limb tremor, shuffling/stooped gait with reduced arm swing | Oscillopsia, dysphagia, bilateral monocular diplopia, pes cavus | Dysequilibrium |  |

| **ID** | **8** | **9** | **10** | **11** | **12** | **13** | **14** | **15** |
| --- | --- | --- | --- | --- | --- | --- | --- | --- |
| **Diagnosis** | *RFC1-*CANVAS | *RFC1-*CANVAS | SCA8 | SCA48 | ATX-*ANO10* (SCAR10) | VCP | SPG7 | 2 variants identified –SCA31 and/or SCA27B |
| **Variant** | *RFC1* biallelic repeat expansion (AAGGG 651/693) | *RFC1* biallelic repeat expansion (AAGGG/ACAGG 1000/2000 repeats) | *ATXN8OS/ATXN8* CTG⋅CAG repeat expansion (94/15 repeats) | Heterozygous *STUB1* in-frame deletion (c.433_435del)  with non-expanded *TBP* (SCA17) (35/36 repeats) | Homozygous *ANO10* splicing variant (c.1163-9A>G) | Heterozygous *VCP* missense variant (c.475C>T, p.Arg159Cys) | Compound heterozygous *SPG7* missense and nonsense variants (c.1045G>A, p.Gly349Ser; c.861dup, p.Asn288Ter) | *BEAN1*  (TAAAA)50(TGGAA)226(TAAAA)120(TGGAA)110(TAAAA)75  *FGF14* GAA expansion (213/201 repeats) |
| **Age (yrs)** | 81-85 | 61-65 | 36-40 | 81-85 | 56-60 | 66-70 | 51-55 | 71-75 |
| **AAO (yrs)** | 76-80  (cough 56-60) | 56-60 | 31-35 | 61-65 | 41-45 | 61-65 | 46-50 | 66-70 |
| **Disease duration (yrs)** | 5 | 3 | 7 | 19 | 15 | 3 | 2 | 4 |
| **Sex** | F | M | F | F | F | M | M | M |
| **FHx** | Sporadic | Sporadic | Sporadic | AD | Sporadic | AD | N/A (adopted) | Sporadic |
| **Primary phenotype** | Ataxia | Ataxia | Ataxia | Ataxia | Ataxia | HSP | Ataxia | Ataxia |
| **Mobility aid** | Nil | Stick | Nil | Wheelchair | 2x sticks | Stick | N/A | Wheelchair |
| **1st symptom** | Vertigo/disequilibrium  (cough onset 20 years earlier) | Collapse after feeling unsteady | Dysarthria | Unsteadiness, balance impairment | Impaired lower limb coordination | Toe curling, stumbling with walking | Gait and balance impairment | Balance impairment |
| **Episodic symptoms** | +** | - | - | - | - | - | - | - |
| **Gait ataxia** | + | + | + | Not testable (wheelchair bound) | + | -  (spastic gait) | + | + |
| **Limb ataxia** | + | + | + | + | + | - | + | + |
| **Dysarthria** | - | + | + | + | + | - | - | + |
| **Nystagmus** | - | DBN | GE | - | GE | - | - | DBN |
| **Head impulse** | N/A | + | N/A | N/A | - | - | N/A | + |
| **Lower limb spasticity** | - | - | - | - | - | + | - | - |
| **Reflexes** | Reduced | Reduced | Normal | Increased | Normal | Increased knee jerks, reduced ankle jerks, upgoing plantar responses | Generally increased reflexes | Normal |
| **Sensory deficit** | + | + | - | - | Focal deficit right hand | + | Deficit dorsal right foot | - |
| **Autonomic dysfunction** | - | + | - | + | - | - | + | - |
| **Cough** | + | - | - | - | - | - | - | - |
| **SARA** | 8.5 | 14 | 8.5 | 30 | 17 | N/A (SPRS 23) | N/A | 18 |
| **MRI/CT abnormalities** | - | Cerebellar atrophy | Cerebellar atrophy | Cerebellar, and brainstem atrophy, generalised age-related cerebral atrophy, basal ganglia calcification | Cerebellar atrophy (more marked in vermis), brainstem and cerebral atrophy | Chronic right cerebellar infarct | Cerebellar atrophy | Cerebellar and cerebral atrophy, basal ganglia mineral deposition, chronic infarct posterior limb left internal capsule |
| **NCS/EMG** | Sensory axonal neuropathy | Sensory axonal neuropathy | N/A | N/A | N/A | N/A | N/A | N/A |
| **Other** | Dysphonia | Oscillopsia, diplopia on lateral gaze | Oscillopsia, right hypertropia, deterioration in symptoms during pregnancies | Memory decline and personality change | Vertigo/dysequilibrium, learning difficulties at school, hearing decline | Lower limb muscle wasting, fasciculations of quadriceps | Slowing of saccades (vertical>horizonal), restricted down-gaze, bilateral ankle clonus | Dysphagia |

AAO: age at onset; DBN: down-beat nystagmus FHx: family history; GE: gaze-evoked nystagmus; HSP: hereditary spastic paraplegia; N/A: not available/unknown *RFC1*-CANVAS: *RFC1*-related cerebellar ataxia, neuropathy, vestibular areflexia syndrome; SCA: spinocerebellar ataxia; SCAR10: autosomal recessive spinocerebellar ataxia-10; SPG7: spastic paraplegia-7; UBN: up-beat nystagmus

*Dysarthria purely episodic with normal speech interictally

**History confounded by diagnosis of benign paroxysmal positional vertigo in setting of episodic vertigo months prior to constant dysequilibrium
