## Supplementary Table 5 for "Targeted long-read sequencing as a single assay improves diagnosis of spastic-ataxia disorders"

**Supplementary table 4. *FGF14* STR expansion lengths on LRS and confirmatory flanking/repeat-primed PCR**

| **ID** | ***FGF14* GAA repeat length** | |
| --- | --- | --- |
|  | **ONT LRS** | **Confirmatory testing – F/RP-PCR** |
| 1 | 296/40 | 302/38 |
| 3 | 321/9 | 330/9 |
| 4 | 274/9 | 274/9 |
| 5 | 387/8 | 414/8 |
| 7 | 338/17 | 340/17 |

F/RP-PCR: flanking and repeat-primed PCR; ONT LRS: Oxford Nanopore Technologies long-read sequencing
